# Pain Among US Adults Before, During, and After the COVID-19 Pandemic: A Repeated Cross-Sectional Study using the 2019-2023 National Health Interview Survey

**DOI:** 10.1101/2024.10.24.24316018

**Authors:** Anna Zajacova, Hanna Grol-Prokopczyk, Richard L. Nahin

## Abstract

**Importance:** Chronic pain (CP) is a major public health problem in the US. The COVID-19 pandemic led to widespread disruptions in the US and it is important to monitor changes in pain during and after the pandemic.

**Objective:** To determine prevalence of chronic pain (CP) and high-impact chronic pain (HICP) before, during, and after the COVID-19 pandemic and identify potential contributing factors.

**Methods:** We analyze a nationally representative sample of 88,469 community-dwelling Americans aged 18 and older from three cross-sectional waves of the National Health Interview Survey before (2019), during (2021), and after (2023) the COVID-19 pandemic.

Year of interview is the exposure. All regression models control for age and sex; fully controlled models include 19 additional covariates (demographics, socioeconomic status, health behaviors, health conditions, mental health, and health insurance type); analyses also explore the role of long COVID. Outcomes are CP and HICP using measures proposed by the US National Pain Strategy; we also present findings for six site-specific pain measures.

**Results:** Between 2019 and 2023, CP and HICP prevalence increased by 18% and 13%, respectively. Specifically, CP prevalence was 20.6% (95%CI: 19.9-21.2%) in 2019, 20.9%(20.3-21.6%) in 2021, and 24.3% (23.7-25.0%) in 2023. HICP prevalence declined from 7.5% (7.1-7.8%) in 2019 to 6.9% (6.6-7.3%) in 2021, before rising sharply to 8.5% (8.1-8.9%) in 2023. The increases occurred in all examined body sites except for tooth/jaw pain, and in all major population groups. Approximately 13% of the increase in CP and HICP was attributable to long COVID.

**Conclusions and Relevance:** Pain among US adults was high before and during the pandemic but has surged substantially since. In 2023, an unprecedented 60 million Americans had chronic pain and 21 million had high-impact chronic pain.

## INTRODUCTION

Chronic pain affects tens of millions of US adults, constituting a public health crisis with profound economic and social consequences. The economic cost of pain---$560-$635 billion annually in 2010 dollars---exceeds the cost of heart disease, cancer, or diabetes.^1^ Pain affects physical, mental, and cognitive health.^2-4^ It also profoundly impacts all other life domains such as employment^5^, family relationships^6^, sexual function,^7^ and sleep,^8^ and contributes significantly to opioid use and misuse, which remain a major public health challenge.^9^ Given pain’s role as a barometer of population health,10 monitoring pain levels in the US is one of the Healthy People 2030 goals.^11^

Chronic pain across all US adult population subgroups increased in the first two decades of the 21st century12-15 but these increases appeared to slow or even reverse by 2020 or 2021.^16,17^ This heartening trend coincided with the start of COVID pandemic, however, when pain researchers warned of potential widespread pain-related consequences of COVID.^18,19^ Pain might increase as a direct consequence of the viral infection;^20,21^ or due to stress, anxiety, depression, and loneliness linked to social distancing,^22,23^ increases in detrimental health behaviors,^24,25^ disrupted health care,^26^ or economic hardships during the pandemic.^27^ Over the past several years, emerging literature has documented pain-related consequences of COVID infection, especially long COVID.^21,28-31^

We are aware of no post-pandemic estimates of chronic pain prevalence in the US, and only one study of pain during the pandemic (in 2021), which found no increase in prevalence, and indeed a slight decrease for high-impact chronic pain.^16^ Moreover, relatively little is known about how social and economic changes during and after the pandemic may have influenced pain prevalence in the US.

Our study provides the first assessment of chronic pain in a nationally-representative sample of US adults before, during, and after the COVID pandemic. We use National Health Interview Survey data and examine multiple dimensions of pain in the adult population and across key demographic subgroups. We examine chronic pain, high-impact chronic pain,^32^ and six location-specific pain types. Finally, we explore potential drivers of the observed changes in pain.

## METHODS

### Data

We analyzed 5-year trends in pain using data from the 2019, 2021, and 2023 National Health Interview Survey (NHIS) harmonized by IPUMS.^33^ The NHIS, approved by the National Center for Health Statistics Research Ethics Review Board, is an ongoing cross-sectional household-based interview survey. It is deemed to be the “best single source for pain surveillance” among US adults, including for the study of pain trends.^34^

The choice of the three waves---2019, 2021, and 2023---is based on NHIS’s sampling and variable collection practices: 2019 is the first year after a major study redesign;35 2023 is the most recent wave available, and pain variables were not collected in 2020 and 2022. In the three waves combined, the NHIS included 90,769 respondents aged 18 and older; 88,469 (97.5%) answered the lead chronic pain question.

### Variables

Two widely-used measures of pain were collected in 2019, 2021, and 2023. Chronic pain (CP) is defined as pain on most days or every day over the past 3 months versus never or some days. High-impact chronic pain (HICP) is CP that also limits activities on most days or every day (versus no CP or CP that limits activities never or only some days).

We also analyze all available site-specific pain questions. Respondents were asked how much pain they experienced during the past 3 months in the following six sites: back; arm, shoulder, or hand; hip, knee, or feet; headache or migraine; abdominal, pelvic, or genital; and jaw or tooth pain. Respondents who said they had “a lot of” pain at a given site (versus none, a little, or ‘somewhere between a little and a lot’) were classified as having significant pain at that site.

The main predictor is the year of interview (2019 as reference, versus 2021, and 2023). Covariates include demographics, socioeconomic status, health behaviors, physical health conditions, and mental health. Variable categorizations and distributions in the population are shown in **Supplemental Table S1**.

The final covariate is long COVID (ever, versus never as reference). Respondents who said they had had COVID were asked “Did you have any symptoms lasting 3 months or longer that you did not have prior to having coronavirus or COVID-19?” This variable is only available in 2023; 2019 was prior to the pandemic and the variable was not collected in 2021.

### Approach

First, we estimated the weighted crude prevalence of CP and HICP and the six site-specific pain measures, in each year, and tested for differences across waves using survey-adjusted Wald tests (**Table 1**). We then visualized the prevalence of CP and HICP in major population groups in each year (**Figure 1**). In addition to the graphical representation, we tested whether the pain increase differed across population subgroups. This was done by estimating regression models of pain with interaction terms between year and population group (**Supplemental Table S2**).

**Table 1.** Prevalence of pain in 2013, 2021, and 2023.

|  | 2019 |  | 2021 |  | 2023 |  | 2021<br>vs<br>2019 | 2023 vs<br>2021 | 2023 vs<br>2019 |
| --- | --- | --- | --- | --- | --- | --- | --- | --- | --- |
|  | N | Prevalence,<br>(95% CI) | N | Prevalence,<br>(95% CI) | N | Prevalence,<br>(95% CI) |  |  |  |
| Sample size | 31,239 |  | 28,687 |  | 28,543 |  |  |  |  |
| Population represented | 250,334,186 |  | 252,461,316 |  | 257,760,200 |  |  |  |  |
| <u>Global pain</u> |  |  |  |  |  |  |  |  |  |
| **Chronic pain** | **50,154,951** | **20.6** (19.9-21.2) | 51,490,798 | 20.9 (20.3-21.6) | **60,713,114** | **24.3** (23.7-25.0) | .358 | <.001 | <.001 |
| **HICP** | **18,218,925** | **7.5** (7.1-7.8) | 17,076,145 | 6.9 (6.6-7.3) | **21,114,932** | **8.5** (8.1-8.9) | .033 | <.001 | <.001 |
| <u>Site-specific pain</u> |  |  |  |  |  |  |  |  |  |
| Back/neck | 27,369,896 | 11.2 (10.8-11.7) | 26,555,370 | 10.8 (10.4-11.2) | 30,992,275 | 12.4 (11.9-12.9) | .132 | <.001 | <.001 |
| Shoulder/arm | 20,774,218 | 8.5 (8.1-9.0) | 19,389,110 | 7.9 (7.5-8.3) | 22,807,394 | 9.2 (8.7-9.6) | .015 | <.001 | .029 |
| Hip/knee/leg | 29,652,413 | 12.2 (11.7-12.7) | 28,200,773 | 11.5 (11.0-12.0) | 32,575,447 | 13.1 (12.6-13.6) | .026 | <.001 | .006 |
| Head/migraine | 11,623,361 | 4.8 (4.5-5.1) | 10,478,580 | 4.3 (4.0-4.6) | 13,832,320 | 5.6 (5.2-5.9) | .010 | <.001 | <.001 |
| Abdominal | 6,162,424 | 2.5 (2.3-2.8) | 5,887,226 | 2.4 (2.2-2.6) | 7,730,317 | 3.1 (2.9-3.4) | .343 | <.001 | <.001 |
| Tooth/jaw | 5,349,077 | 2.2 (2.0-2.4) | 5,277,190 | 2.2 (1.9-2.4) | 5,534,036 | 2.2 (2.0-2.4) | .736 | .602 | .871 |

Weighted prevalence; confidence estimate calculations adjusted for complex sampling design. Weights for tests across three years are adjusted for data pooling. P-values are from survey-adjusted Wald tests for differences across years. Source: National Health Interview Survey, Adults 18 and older. HICP=high-impact chronic pain.

**Figure 1.**
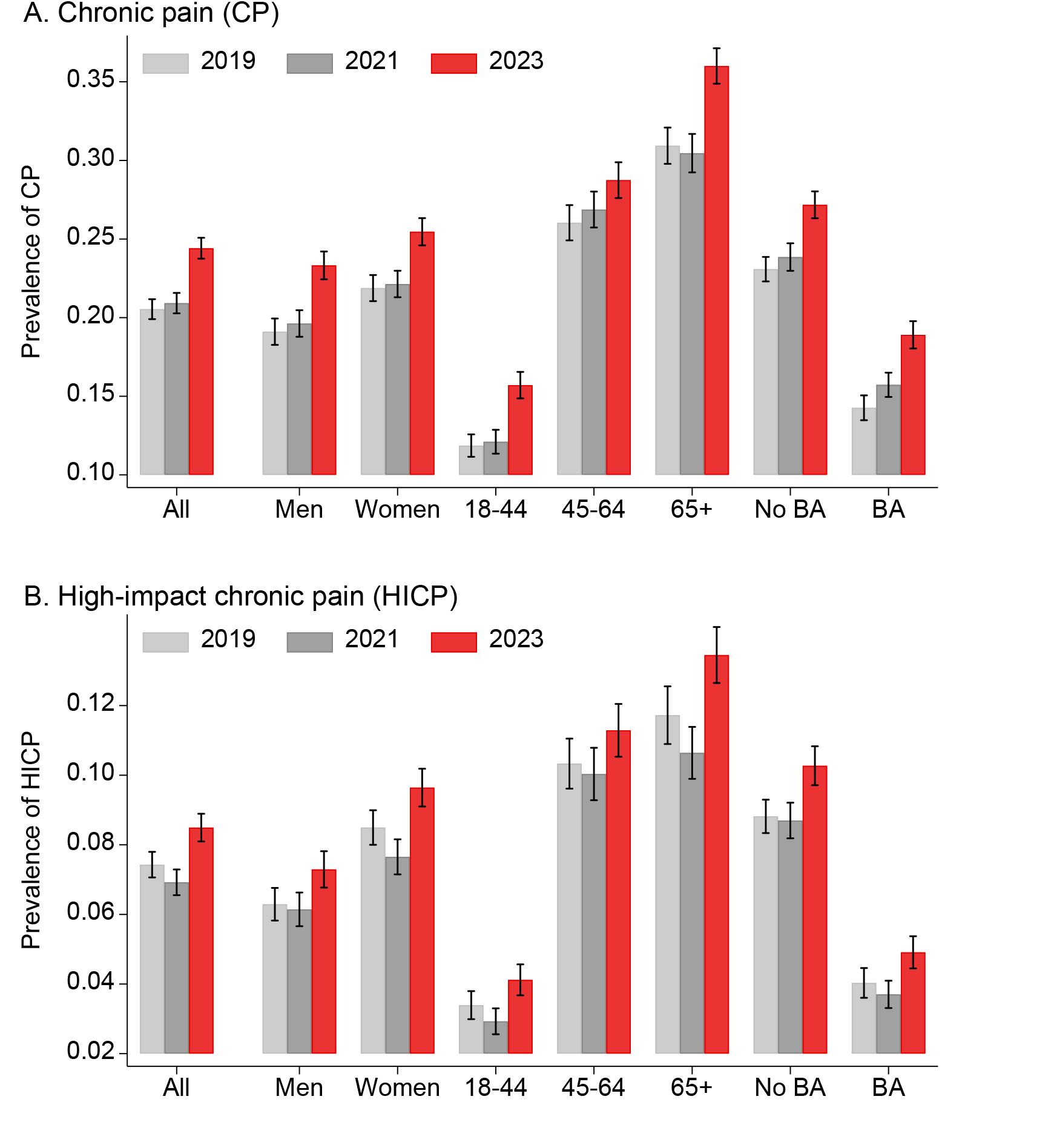
Chronic pain in 2019, 2021, and 2023, US adults age 18+. Note: Weighted prevalence estimates and 95% confidence intervals adjusted for complex sampling design. Population in aggregate (all), by sex (men and women), by age (18-44, 45-64, and 65+), and by education (no college degree [no BA] and has college degree [BA])

Second, we examined the covariates of pain change over time using regression-based approaches. We estimated a series of regression models of all pain outcomes as a function of year, net of different covariate groups: Model 1 was bivariate, including only the survey year; Model 2 controlled for all covariates except long COVID; and Model 3 added long COVID (**Table 2**). We used modified Poisson models, which are appropriate for dichotomous outcomes and increasingly preferred over logistic models because their exponentiated coefficients are interpretable as prevalence ratios rather than the less intuitive odds ratios.^36^ Table 2 also shows the percent of pain change between 2019 and 2023 that was due to long COVID, estimated using the Karlson-Breen-Holm (KHB) method^37^ for comparing coefficient effects across nonlinear models.

**Table 2.**
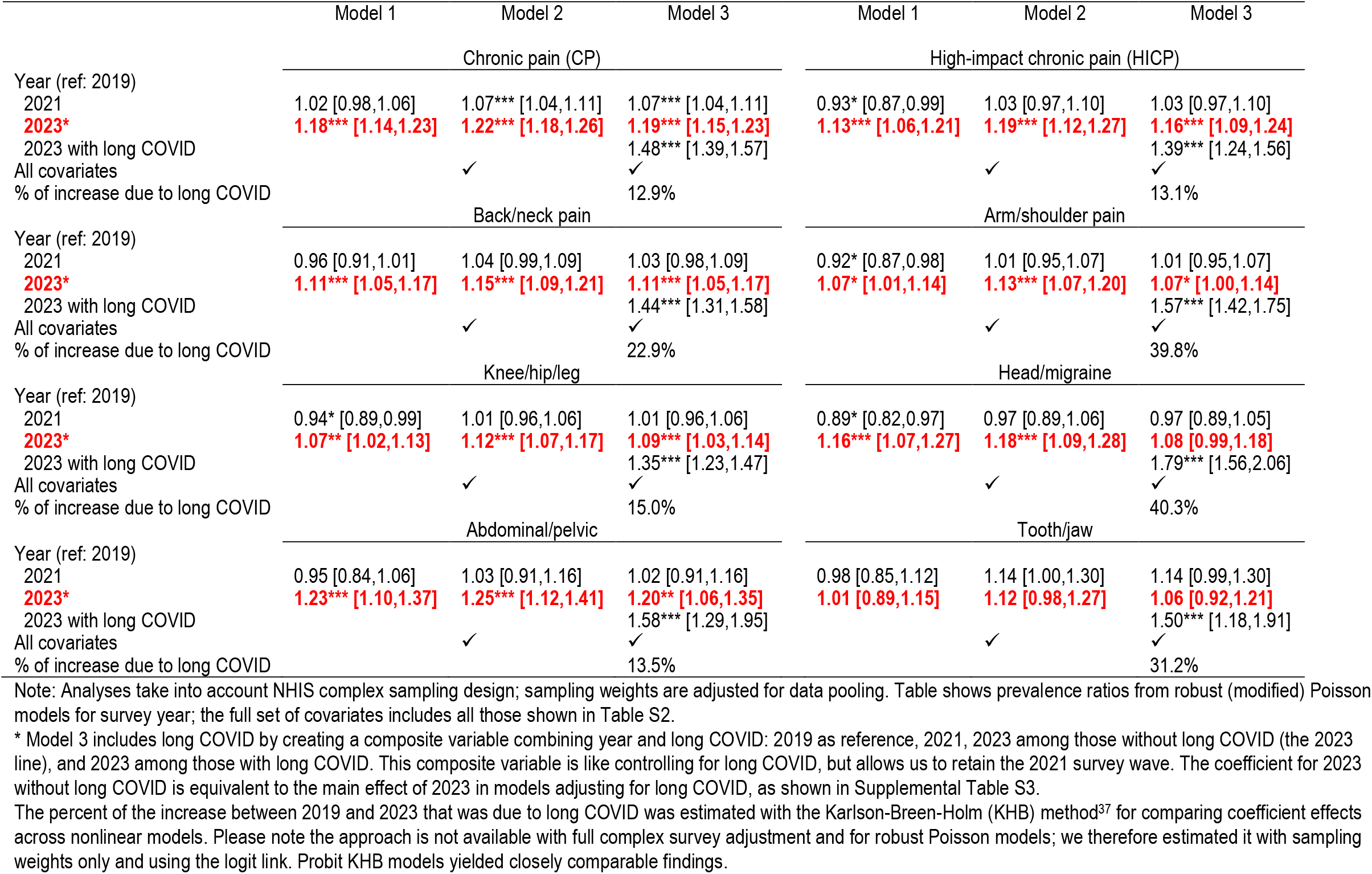
Prevalence ratios for CP, HICP, and site-specific pain in 2021 and 2023 relative to 2019 under different model specifications.

We conducted extensive supplemental analyses to assess the robustness of our conclusions to different variable and model specifications. The regression models were re-estimated with logit, probit, and linear probability models. The findings were comparable to those shown. We also incorporated long COVID in the regression models in three different ways due to its collinearity with 2019 and absence in 2021 (**Supplemental Table S3**). Again, the findings were robust to these different model specifications. Finally, we closely examined the role of long COVID in shaping pain prevalence in 2023. First, we estimated the association between long COVID and pain net of covariates; this set of models yielded estimates of the long COVID-pain association at the individual level (**Supplemental Table S4**). Then we analyzed the role of long COVID at the population level. We calculated the counterfactual (hypothetical) prevalence of pain under two scenarios: if no one had long COVID, and if everyone in the population had long COVID. These were obtained by calculating the predicted probabilities of the outcome (pain) for each individual using their actual covariate values, but assigning the values of 0 or 1, respectively, for long COVID. We then averaged these probabilities over all observations. The comparison of the first scenario and actual observed prevalence was also used to calculate the population-attributable fraction38 of CP and HICP due to long COVID (**Supplemental Table S5**).

## RESULTS

**Table 1** shows the weighted crude prevalence of pain in each year. The prevalence of CP was 20.6% (95%CI 19.9-21.2%) in 2019 and remained statistically unchanged at 20.9% (95%CI 20.3-21.6%) in 2021 but increased significantly to 24.3% (95%CI 23.7-25.0%) in 2023. HICP prevalence was 7.5% (95%CI 7.1-7.9%) in 2019. It declined significantly to 6.9% (95%CI 6.6-7.3%) in 2021 but increased to 8.5% (95%CI 8.1-8.9%) in 2023.

Table 1 also summarizes the prevalence of the six site-specific pain measures. Tooth/jaw pain remained stable across all waves. All other sites remained stable or declined significantly in 2021 versus 2019 but then increased significantly by 2023, so that five of the six sites (back/neck, shoulder/arm, hip/knee/leg, headache/migraine, and abdominal/pelvic) had higher prevalence in 2023 than in both 2021 and 2019. For example, the highest-prevalence site— hip/knee/leg pain—declined significantly from 12.2% (95%CI 11.7-12.7%) in 2019 to 11.5% (95%CI 11.0-12.0%) in 2021, then increased to 13.1% (95%CI 12.6-13.6%) in 2023.

**Figure 1** visualizes the patterns in CP (Plot A) and HICP (Plot B) in the full sample, and also shows the prevalences by age, sex, and educational attainment. The graphs show that the 2023 increase was widespread across major population groups: CP and HICP stayed relatively stable in 2021 compared with 2019, but increased sharply in 2023. Results by race/ethnicity are in **Supplemental Figure S1** and show similar temporal patterns.

In addition, we also tested whether the 2023 increases relative to 2019 were comparable by age, sex, educational attainment, and race/ethnicity. The findings are in **Supplemental Table S2**. The HICP increases were statistically equivalent across all population groups. CP increases were also comparable for men and women, as well as for Black and Hispanic, as compared with white, adults. However, CP increased significantly more for younger people, Asian Americans, and college graduates as compared with older adults, whites, and those without a college degree, respectively.

The second part of the analysis explored explanations for the changes in pain prevalence. **Supplemental Table S1** shows that the distribution of population characteristics relevant to pain prevalence in each year. Demographics (age, sex, region, race/ethnicity, foreign-born) did not change significantly and neither did the prevalence of physical chronic conditions (angina, arthritis, cancer, CHD, diabetes, myocardial infarction, or hypertension). However, the distributions of socioeconomic factors, mental health, health behaviors, and health insurance changed significantly between 2019 and 2023: Education and family income increased, and a smaller proportion were uninsured or had difficulty paying medical bills. Smoking prevalence decreased but the prevalence of obesity increased significantly, from 31.2% in 2019 to 33.3% in 2023. The prevalence of elevated depressive symptoms and anxiety increased significantly as well. The final covariate is long COVID. In 2023, 8.3% of the population reported having had long COVID. This includes those who had long COVID previously but recovered prior to the interview (4.8%), those with current long COVID (3.5%), and those with current high-impact long COVID (0.7%).

**Table 2** includes these characteristics in regression models of CP, HICP, and site-specific pain. Model 1 is unadjusted; Model 2 includes all covariates except long COVID; and Model 3 adds long COVID. Regarding CP, the unadjusted prevalence was unchanged in 2021 versus 2019 but increased 18% in 2023 (PR=1.18, 95%CI 1.14,1.23). Net of the covariates added in Model 2, the 2023 prevalence remained significantly higher (PR=1.22, 95%CI 1.18,1.26). In Model 3, we added a composite year/long-COVID variable with four categories: 2019 (reference), 2021, 2023 *without* long COVID (yielding year effect estimates identical to those in models controlling for long COVID), and 2023 *with* long COVID. In this Model, the 2023 prevalence of CP net of long COVID was 19% higher compared with 2019 (PR=1.19, 95%CI 1.15,1.23). The table provides two perspectives on the role of long COVID in CP trends. First, among adults who reported in 2023 that they ever had long COVID, pain was 48% higher than in the 2019 population (PR=1.48, 95%CI 1.39,1.57). Second, the proportion of the population-level increase in chronic pain attributable to long COVID, obtained via the KHB decomposition of estimates in Models 2 and 3, was 12.9%.

Table 2 also shows the same series of models for HICP and site-specific pain. Consistently for all these outcomes, the data show 1) no significant increase in prevalence in 2021 compared with 2019; 2) a significant increase in 2023 compared with 2019 (except for tooth/jaw pain); 3) net of all sociodemographic covariates in Model 2, the same patterns hold; in fact, the prevalence ratios for 2023 are even larger than in Model 1; and 4) long COVID explains part of the increase between 2019 and 2023. Specifically, the percent increase in pain prevalence due to long COVID ranges from approximately 13-15% for HICP, abdominal, and knee/hip/leg pain up to 40% for headache/migraine and arm/shoulder pain. (Regarding finding #3: While comparing coefficients across nonlinear models is potentially problematic, sensitivity analyses using linear probability models, also appropriate for dichotomous outcomes,39 revealed the same suppressor effects of the covariates.)

Table 2 only shows the coefficients of the key predictors for parsimony; full results of Model 3 for CP and HICP are shown in **Supplemental Table S3**. This table also shows the equivalence of three ways of including long COVID in the models: using the composite variable as in Table 2; including long COVID as a covariate, and excluding adults with long COVID.

Finally, we offer two additional perspectives on the role of long COVID in pain prevalence by analyzing only 2023 data. **Supplemental Table S4** shows the results of regression models of CP and HICP as a function of long COVID, disaggregating this variable into long COVID in the past, current long COVID, and current high-impact long COVID. The results show a dose-response relationship between these COVID variables and pain, confirming the strong association between long COVID and pain at the individual level.

**Supplemental Table S5** returns to the population level and calculates the population-attributable fraction of CP and HICP due to long COVID in 2023. Adjusting only for age and sex, about 5-7% of pain in the population is due to long COVID, a nontrivial but not a decisive proportion. The proportion-attributable fraction is below 3% in fully-adjusted models.

Counterfactual simulations (hypothetical estimates) show that if nobody in the population had long COVID, CP and HICP in 2023 would have been 23.0% (95% CI 22.4, 23.6) and 7.8% (95% CI 7.5, 8.2)—still higher than in 2019 but slightly less than the actually observed prevalences.

## DISCUSSION

Pain surged dramatically among US adults after the COVID pandemic. From 2019 to 2023, chronic pain and high-impact chronic pain prevalence increased by 18% and 13%, respectively. The widely cited 20% population prevalence of CP^16,40,41^ appears outdated; our updated estimate from 2023 is 24.3%. This increase represents more than 10 million additional individuals experiencing chronic pain in 2023 compared with just five years prior, bringing the total to over 60 million adults. Similarly, high-impact chronic pain, the most debilitating form of pain,^32^ affected 8.5% of adults in 2023, up from 7.5% in 2019 and only 6.9% during the pandemic in 2021. This new estimate translates to over 21 million Americans experiencing high-impact pain in 2023.

The increase is not restricted to specific demographics or pain sites: It is evident in most population subgroups and in all examined pain sites except tooth/jaw pain. We found similar relative increases in HICP for younger and older adults, men and women, college graduates and people without college diplomas, and across all race/ethnic groups. The increases in CP were often similar across groups as well, but groups with lower prevalences before the pandemic (younger adults, Asian Americans, and college graduates) experienced significantly steeper increases than older adults, whites, and people without college diplomas, suggesting some convergence in age-related, racial/ethnic, and educational disparities in pain.

The 2023 increase was not explained by changes in the population distribution of important sociodemographic and health covariates including age, education, economic well-being, smoking and obesity, physical conditions, and depressive and anxiety symptoms. In fact, if the population distribution of these characteristics had remained at 2019 levels, the increase in pain might have been even greater. However, this interpretation warrants caution given the cross-sectional and observational nature of our data.

One important factor is long COVID. At the individual level, long COVID is powerfully associated with pain,^29,42^ a relationship we confirmed in our analyses of 2023 data: adults with long COVID are significantly more likely to report both chronic and high-impact pain. However, long COVID explains only part of the 2019-to-2023 increase in pain prevalence. For chronic and high-impact chronic pain, long COVID explained about 13% of the increase. For headache/migraine and arm/shoulder pain, the role of long COVID was larger: it explained about 40% of the increases.

Our findings suggest that broader social and systemic factors beyond our measured covariates are driving the post-pandemic pain surge. These may include social connections, a powerful correlate of chronic pain.^43-45^ Social isolation, loneliness, and heightened levels of stress and anxiety, as well as disrupted health care access—all of which were exacerbated by the pandemic—should be explored in future research. Another possibility is that reporting tendencies changed over time, although it is not obvious what factors would cause a nation-wide change in pain reporting over a period of just two years.

Limitations of our study include its use of cross-sectional data. Longitudinal data could allow causal analyses that might better identify the reasons for the pain increase. In addition, NHIS survey collection changed from mostly in-person in 2019 to mostly by phone in 2021 to a mixed mode in 2023; it is unclear how this might have influenced 6findings. We also caution that NHIS only includes non-institutionalized adults, so changes in institutionalization might contribute to observed patterns. Finally, all covariates were self-reported. This is potentially problematic, especially with respect to reports of long COVID. Given the complexity and recent emergence of this condition, over- and/or under-reporting are possible; this may lead to mis-estimation of its role in the rising prevalence of pain.

Despite limitations, this analysis is the first comprehensive report on the escalation of chronic and high-impact pain after the pandemic in the US adult population. Our findings should be replicated with other nationally-representative data sources, and a broader set of potential explanatory factors should be employed to explain the post-pandemic trends. We found that chronic pain, already a widespread health problem, reached an all-time high in prevalence in the post-pandemic era, necessitating urgent attention and interventions to address and alleviate this growing health crisis. Pain serves as a sensitive barometer of population health10 and has profound economic, social, and health consequences. It is thus critical to address the rise in chronic pain.

## Data Availability

All data used in the present study are available publicly online at https://nhis.ipums.org/nhis/. All Stata code is available on request to the authors.

