## Supplement for "Pain Among US Adults Before, During, and After the COVID-19 Pandemic: A Repeated Cross-Sectional Study using the 2019-2023 National Health Interview Survey"

### SUPPLEMENTAL MATERIAL

**Supplemental Table S1. Sample characteristics in 2019, 2021, and 2023**

| | Survey year | | | |
|---|---|---|---|---|
| | 2019 | 2021 | 2023 | Test |
| Population size | 250,334,186 | 252,461,316 | 257,760,200 | |
| Age (mean, s.d.) | 47.7 (18.3) | 48.1 (18.4) | 48.1 (18.5) | 0.081 |
| Female | 129,412,107 (51.7%) | 130,389,168 (51.7%) | 131,939,225 (51.2%) | 0.539 |
| Region of residence | | | | 0.954 |
| Northeast | 44,492,120 (17.8%) | 44,060,428 (17.5%) | 44,992,836 (17.5%) | |
| Midwest | 52,661,442 (21.0%) | 52,520,602 (20.8%) | 53,077,517 (20.6%) | |
| South | 94,302,024 (37.7%) | 95,794,273 (37.9%) | 98,869,532 (38.4%) | |
| West | 58,878,600 (23.5%) | 60,086,012 (23.8%) | 60,820,315 (23.6%) | |
| Race/ethnicity | | | | 0.260 |
| White | 158,304,655 (63.2%) | 158,598,869 (62.8%) | 159,522,980 (61.9%) | |
| Black | 29,378,731 (11.7%) | 29,430,773 (11.7%) | 30,364,132 (11.8%) | |
| Hispanic | 41,439,638 (16.6%) | 42,761,449 (16.9%) | 45,261,461 (17.6%) | |
| Asian | 14,703,362 (5.9%) | 14,937,005 (5.9%) | 16,187,251 (6.3%) | |
| Other | 6,507,800 (2.6%) | 6,733,220 (2.7%) | 6,424,376 (2.5%) | |
| Foreign-born | 45,162,051 (18.6%) | 44,769,769 (18.3%) | 46,729,314 (19.0%) | 0.393 |
| Marital status | | | | 0.004 |
| Married/cohabiting | 149,086,612 (61.3%) | 147,409,597 (60.4%) | 148,533,425 (60.3%) | |
| Previously married | 39,293,506 (16.2%) | 38,461,741 (15.8%) | 38,497,247 (15.6%) | |
| Never married | 54,834,692 (22.5%) | 58,306,151 (23.9%) | 59,350,602 (24.1%) | |
| Education | | | | <0.001 |
| LHS/GED | 36,791,321 (14.8%) | 29,671,063 (11.8%) | 32,861,727 (12.8%) | |
| HS | 62,361,506 (25.1%) | 65,324,595 (26.0%) | 63,733,613 (24.9%) | |
| SC/AA | 77,352,019 (31.1%) | 66,749,842 (26.6%) | 74,926,357 (29.2%) | |
| BA+ | 72,039,247 (29.0%) | 89,168,590 (35.5%) | 84,705,791 (33.1%) | |
| Food insecurity | 20,798,082 (8.3%) | 14,229,124 (5.6%) | 22,119,891 (8.6%) | <0.001 |
| Difficult to pay med. bills | 34,235,025 (13.8%) | 26,443,750 (10.5%) | 27,361,167 (10.7%) | <0.001 |
| Family income | | | | <0.001 |
| Below poverty line (PL) | 28,088,434 (11.2%) | 24,953,123 (9.9%) | 25,919,804 (10.1%) | |
| 1-1.99 times PL | 46,787,583 (18.7%) | 44,286,036 (17.5%) | 46,479,277 (18.0%) | |
| 2-4.99 times PL | 103,315,302 (41.3%) | 104,206,825 (41.3%) | 104,206,062 (40.4%) | |
| 5 or more times PL | 72,142,867 (28.8%) | 79,015,331 (31.3%) | 81,155,057 (31.5%) | |
| Body mass index | | | | <0.001 |
| BMI < 18.5 | 4,012,552 (1.6%) | 4,326,250 (1.7%) | 4,059,011 (1.6%) | |
| 18.5<=BMI<25 | 78,561,434 (31.4%) | 77,314,310 (30.6%) | 77,284,221 (30.0%) | |
| 25<=BMI<30 | 82,616,122 (33.0%) | 84,013,937 (33.3%) | 85,755,233 (33.3%) | |
| BMI>=30 | 78,209,385 (31.2%) | 81,015,576 (32.1%) | 85,511,666 (33.2%) | |
| Unknown | 6,934,693 (2.8%) | 5,791,243 (2.3%) | 5,150,069 (2.0%) | |
| Smoking status | | | | <0.001 |
| Current | 34,015,663 (14.0%) | 28,289,579 (11.6%) | 26,952,716 (10.8%) | |
| Former | 54,828,260 (22.5%) | 56,266,805 (23.0%) | 55,887,495 (22.5%) | |

| | | | | |
|---|---|---|---|---|
| Never | 154,376,037 (63.5%) | 160,281,836 (65.5%) | 165,699,129 (66.7%) | |
| Angina | 4,209,203 (1.7%) | 3,805,363 (1.5%) | 4,112,172 (1.6%) | 0.295 |
| Arthritis | 53,534,488 (21.4%) | 53,685,652 (21.3%) | 54,664,027 (21.2%) | 0.886 |
| Cancer | 23,727,697 (9.5%) | 24,655,468 (9.8%) | 24,961,410 (9.7%) | 0.550 |
| CHD | 11,513,251 (4.6%) | 12,400,324 (4.9%) | 12,403,444 (4.8%) | 0.214 |
| Diabetes | 23,419,131 (9.4%) | 24,254,223 (9.6%) | 25,264,113 (9.8%) | 0.253 |
| Heart attack | 7,863,199 (3.1%) | 7,679,071 (3.0%) | 7,780,823 (3.0%) | 0.707 |
| Hypertension | 79,155,739 (31.7%) | 79,386,312 (31.5%) | 83,015,797 (32.3%) | 0.220 |
| High depression symptoms | 23,577,423 (9.6%) | 24,667,027 (10.0%) | 26,367,464 (10.5%) | 0.014 |
| High anxiety symptoms | 31,104,614 (12.7%) | 31,754,918 (12.9%) | 35,067,142 (14.0%) | <0.001 |
| Health insurance | | | | <0.001 |
| Private | 159,133,580 (63.8%) | 162,229,570 (64.5%) | 161,987,083 (63.2%) | |
| None | 28,748,196 (11.5%) | 24,751,534 (9.8%) | 21,505,121 (8.4%) | |
| Public | 61,350,733 (24.6%) | 64,348,414 (25.6%) | 72,953,116 (28.4%) | |
| Long COVID | | | 21,290,971 (8.3%) | |
| Detailed COVID breakdown | | | | |
| No COVID | | | 116,034,322 (45.0%) | |
| COVID but not long | | | 120,434,907 (46.7%) | |
| Long but not now | | | 12,130,433 (4.7%) | |
| Long now but not high-impact | | | 7,265,610 (2.8%) | |
| HILC | | | 1,894,928 (0.7%) | |

Weighted and adjusted for complex sampling design. Weights for tests across three years are adjusted for data pooling. P-values are from survey-adjusted Wald tests for difference across years. Source: National Health Interview Survey, adults 18 and older. HICP=high-impact chronic pain; HILC=high-impact long COVID.

**Supplemental Table S2. Models of CP and HICP comparing the 2023 vs 2019 increase by age, sex, race/ethnicity, and education.**

| | **Chronic pain** | | | | | | | | **High-impact chronic pain** | | | | | | | |
|---|---|---|---|---|---|---|---|---|---|---|---|---|---|---|---|---|
| 2023 vs 2019 | 1.38*** | 1.20*** | 1.17*** | 1.16*** | 1.42*** | 1.23*** | 1.20*** | 1.19*** | 1.21 | 1.13** | 1.12** | 1.14*** | 1.22 | 1.17** | 1.17*** | 1.18*** |
| Age (continuous) | 1.02*** | 1.02*** | 1.02*** | 1.02*** | 1.01*** | 1.01*** | 1.01*** | 1.01*** | 1.03*** | 1.03*** | 1.03*** | 1.03*** | 1.01*** | 1.01*** | 1.01*** | 1.01*** |
| Female | 1.08*** | 1.10*** | 1.08*** | 1.07*** | 0.98 | 0.99 | 0.98 | 0.98 | 1.27*** | 1.28*** | 1.27*** | 1.25*** | 1.08* | 1.07 | 1.08* | 1.08* |
| Race (White as ref) | | | | | | | | | | | | | | | | |
| Black | | | 0.90* | | 0.85*** | 0.85*** | 0.86*** | 0.85*** | | | 1.00 | | 0.87** | 0.87** | 0.86* | 0.87** |
| Hispanic | | | 0.65*** | | 0.79*** | 0.79*** | 0.74*** | 0.79*** | | | 0.79** | | 0.91 | 0.91 | 0.88 | 0.91 |
| Asian | | | 0.32*** | | 0.67*** | 0.67*** | 0.56*** | 0.67*** | | | 0.30*** | | 0.59*** | 0.59*** | 0.56** | 0.59*** |
| Other | | | 1.27*** | | 0.99 | 0.99 | 1.01 | 0.99 | | | 1.38* | | 0.98 | 0.98 | 0.97 | 0.98 |
| Education (BA or more) | | | | 0.62*** | | | | 0.83*** | | | | 0.46*** | | | | 0.79*** |
| **Interaction effects** | | | | | | | | | | | | | | | | |
| 2023 by age | 1.00** | | | | 1.00** | | | | 1.00 | | | | 1.00 | | | |
| 2023 by female | | 0.96 | | | | 0.98 | | | | 0.98 | | | | 1.01 | | |
| 2023 by Black | | | 0.94 | | | | 0.96 | | | | 1.01 | | | | 1.02 | |
| 2023 by Hispanic | | | 1.10 | | | | 1.11 | | | | 1.09 | | | | 1.05 | |
| 2023 by Asian | | | 1.44* | | | | 1.35* | | | | 1.06 | | | | 1.07 | |
| 2023 by Other | | | 0.90 | | | | 0.97 | | | | 0.92 | | | | 1.02 | |
| 2023 by BA or more | | | | 1.14** | | | | 1.09* | | | | 1.06 | | | | 1.00 |
| Covariates | | | | | ✓ | ✓ | ✓ | ✓ | | | | | ✓ | ✓ | ✓ | ✓ |

Note: analyses take into account NHIS complex sampling design; sampling weights are adjusted for data pooling. Prevalence ratios from robust (modified) Poisson models shown. Models include only 2019 and 2023 waves for parsimony.

**Supplemental Table S3. Prevalence ratios for CP and HICP in 2021 and 2023 relative to 2019 under different model specifications**

| | Chronic pain (CP) | | | High-impact chronic pain (HICP) | | |
|---|---|---|---|---|---|---|
| | Model 1 | Model 2 | Model 3 | Model 1 | Model 2 | Model 3 |
| **Year (2019)** | | | | | | |
| 2021 | 1.07*** | 1.07*** | 1.07*** | 1.03 | 1.03 | 1.03 |
| **2023*** | **1.19*** ** | **1.19*** ** | **1.19*** ** | **1.16*** ** | **1.16*** ** | **1.17*** ** |
| **2023 with long COVID** | **1.48*** ** | | | **1.39*** ** | | |
| **Had long COVID** | | **1.24*** ** | | | **1.19** ** | |
| Age | 1.01*** | 1.01*** | 1.01*** | 1.01*** | 1.01*** | 1.01*** |
| Female | 0.97 | 0.97 | 0.98 | 1.04 | 1.04 | 1.04 |
| Region (Northeast) | | | | | | |
| Midwest | 1.09** | 1.09** | 1.09** | 1.06 | 1.06 | 1.06 |
| South | 1.07** | 1.07** | 1.08** | 1.11* | 1.11* | 1.12* |
| West | 1.21*** | 1.21*** | 1.21*** | 1.29*** | 1.29*** | 1.28*** |
| Race (White) | | | | | | |
| Black | 0.84*** | 0.84*** | 0.84*** | 0.89** | 0.89** | 0.88** |
| Hispanic | 0.81*** | 0.81*** | 0.80*** | 0.89* | 0.89* | 0.89* |
| Asian American | 0.64*** | 0.64*** | 0.63*** | 0.55*** | 0.55*** | 0.56*** |
| Other | 0.98 | 0.98 | 0.99 | 1.03 | 1.03 | 1.05 |
| Foreign-born (US-born) | 0.82*** | 0.82*** | 0.81*** | 0.87* | 0.87* | 0.87** |
| Marital status (married) | | | | | | |
| Previously married | 0.97* | 0.97* | 0.97 | 0.96 | 0.96 | 0.96 |
| Never married | 0.82*** | 0.82*** | 0.82*** | 0.84*** | 0.84*** | 0.83*** |
| Education (BA+) | | | | | | |
| Less than high school | 1.09*** | 1.09*** | 1.09** | 1.23*** | 1.23*** | 1.21*** |
| High school diploma | 1.07** | 1.07** | 1.07** | 1.20*** | 1.20*** | 1.19*** |
| Some college or AA | 1.14*** | 1.14*** | 1.15*** | 1.26*** | 1.26*** | 1.27*** |
| Food insecure | 1.18*** | 1.18*** | 1.19*** | 1.38*** | 1.38*** | 1.43*** |
| Diff. paying medical bills | 1.31*** | 1.31*** | 1.32*** | 1.50*** | 1.50*** | 1.52*** |
| Family income (5 times PL) | | | | | | |
| Below poverty line (PL) | 1.19*** | 1.19*** | 1.20*** | 1.63*** | 1.63*** | 1.67*** |
| 1-1.99 times PL | 1.10*** | 1.10*** | 1.10*** | 1.32*** | 1.32*** | 1.33*** |
| 2-4.99 times PL | 1.05** | 1.05** | 1.05* | 1.19*** | 1.19*** | 1.20*** |
| BMI (normal weight) | | | | | | |
| Underweight | 1.07 | 1.07 | 1.07 | 1.38*** | 1.38*** | 1.38*** |
| Overweight | 1.11*** | 1.11*** | 1.11*** | 1.03 | 1.03 | 1.03 |
| Obese | 1.29*** | 1.29*** | 1.30*** | 1.27*** | 1.27*** | 1.28*** |
| Unknown BMI | 1.11* | 1.11* | 1.09 | 1.15 | 1.15 | 1.16 |
| Smoking (Never) | | | | | | |
| Current | 1.35*** | 1.35*** | 1.36*** | 1.34*** | 1.34*** | 1.36*** |
| Former | 1.21*** | 1.21*** | 1.22*** | 1.21*** | 1.21*** | 1.21*** |
| Angina | 1.04 | 1.04 | 1.05 | 1.12 | 1.12 | 1.13* |
| Arthritis | 2.47*** | 2.47*** | 2.49*** | 3.01*** | 3.01*** | 2.99*** |
| Cancer | 1.06*** | 1.06*** | 1.07*** | 1.08* | 1.08* | 1.09** |
| CHD | 1.05* | 1.05* | 1.06* | 1.09 | 1.09 | 1.10* |
| Diabetes | 1.14*** | 1.14*** | 1.14*** | 1.23*** | 1.23*** | 1.22*** |
| Heart attack | 0.99 | 0.99 | 0.98 | 1.07 | 1.07 | 1.06 |
| Hypertension | 1.13*** | 1.13*** | 1.14*** | 1.27*** | 1.27*** | 1.26*** |
| High depressive symptoms | 1.41*** | 1.41*** | 1.43*** | 1.92*** | 1.92*** | 1.95*** |
| High anxiety symptoms | 1.48*** | 1.48*** | 1.48*** | 1.66*** | 1.66*** | 1.65*** |
| Health insurance (private) | | | | | | |

| | | | | | | |
|---|---|---|---|---|---|---|
| No insurance | 1.01 | 1.01 | 1.02 | 0.88 | 0.88 | 0.90 |
| Public | 1.10*** | 1.10*** | 1.10*** | 1.33*** | 1.33*** | 1.34*** |

*In Model 1, this coefficient is the relative prevalence of the outcome in 2023 among those without long COVID, relative to 2019.
Note: analyses take into account NHIS complex sampling design; sampling weights are adjusted for data pooling. Prevalence ratios from robust (modified) Poisson models. Model 3 excludes respondents who reported currently or previously having long COVID.

| Supplemental Table S4. Prevalence ratios for CP and HICP as a function of COVID, 2023 data only. | | | | | |
|---|---|---|---|---|---|
| | | Chronic pain (CP) | | High-impact chronic pain (HICP) | |
| | 2023 COVID status | Model 1 | Model 2 | Model 1 | Model 2 |
| COVID status (ref: never had) | 44.7% | | | | |
| Had COVID | 46.9% | 1.09*** | 1.17*** | 0.95 | 1.10* |
| Had long COVID | 4.7% | 1.64*** | 1.45*** | 1.47*** | 1.23* |
| Has long COVID now | 2.9% | 1.92*** | 1.42*** | 1.91*** | 1.24* |
| Has high-impact long COVID | 0.7% | 2.78*** | 1.32*** | 5.01*** | 1.71*** |
| Covariates | | | | | |
| Age | | 1.02*** | 1.01*** | 1.03*** | 1.01*** |
| Female | | 1.06*** | 0.97 | 1.24*** | 1.07* |
| All covariates | | | ✓ | | ✓ |

Note: analyses take into account NHIS complex sampling design; sampling weights are adjusted for data pooling. Prevalence ratios from robust (modified) Poisson models. The full set of covariates includes all those shown in Table S2.

| Supplemental Table S5. Long-COVID counterfactual scenarios and population-attributable fraction of CP and HICP in 2023. | | |
|---|---|---|
| | CP | HICP |
| Percent of pain attributable to long COVID | | |
| Age and sex-adjusted PAF | 5.5% (4.7%-6.3%) | 7.3% (5.7%-8.9%) |
| Fully-adjusted PAF | 2.7% (2.0%-3.4%) | 2.6% (1.0%-4.1%) |
| Observed and hypothetical prevalence values | | |
| Actual observed prevalence (as in Table 1) | 24.3% (23.7%-25.0%) | 8.5% (8.1%-8.9%) |
| Age and sex-adjusted prevalence | | |
| If nobody had long COVID (hypothetical) | 23.0% (22.4%-23.6%) | 7.8% (7.5%-8.2%) |
| If everybody had long COVID (hypothetical) | 39.1% (36.9%-41.3%) | 15.5% (13.8%-17.1%) |
| Fully-adjusted counterfactuals | | |
| If nobody had long COVID (hypothetical) | 23.7% (23.1%-24.2%) | 8.2% (7.8%-8.5%) |
| If everybody had long COVID (hypothetical) | 30.8% (28.9%-32.7%) | 10.1% (9.0%-11.3%) |

Weighted. PAF=population-attributable fraction of outcome (CP and HICP, respectively) due to long COVID. PAF is calculated as (observed prevalence – hypothetical prevalence with no long COVID)/observed prevalence × 100. The counterfactual (hypothetical) values are calculated by: (1) estimating a logistic regression model of the pain outcome; (2) calculating the predicted probability of pain for each individual, using their actual covariate values but assigning long COVID=0 to everyone; and (3) averaging these probabilities over all observations.

**Supplemental Figure S1. Chronic pain in 2019, 2021, and 2023, US adults age 18+, by race/ethnicity.**

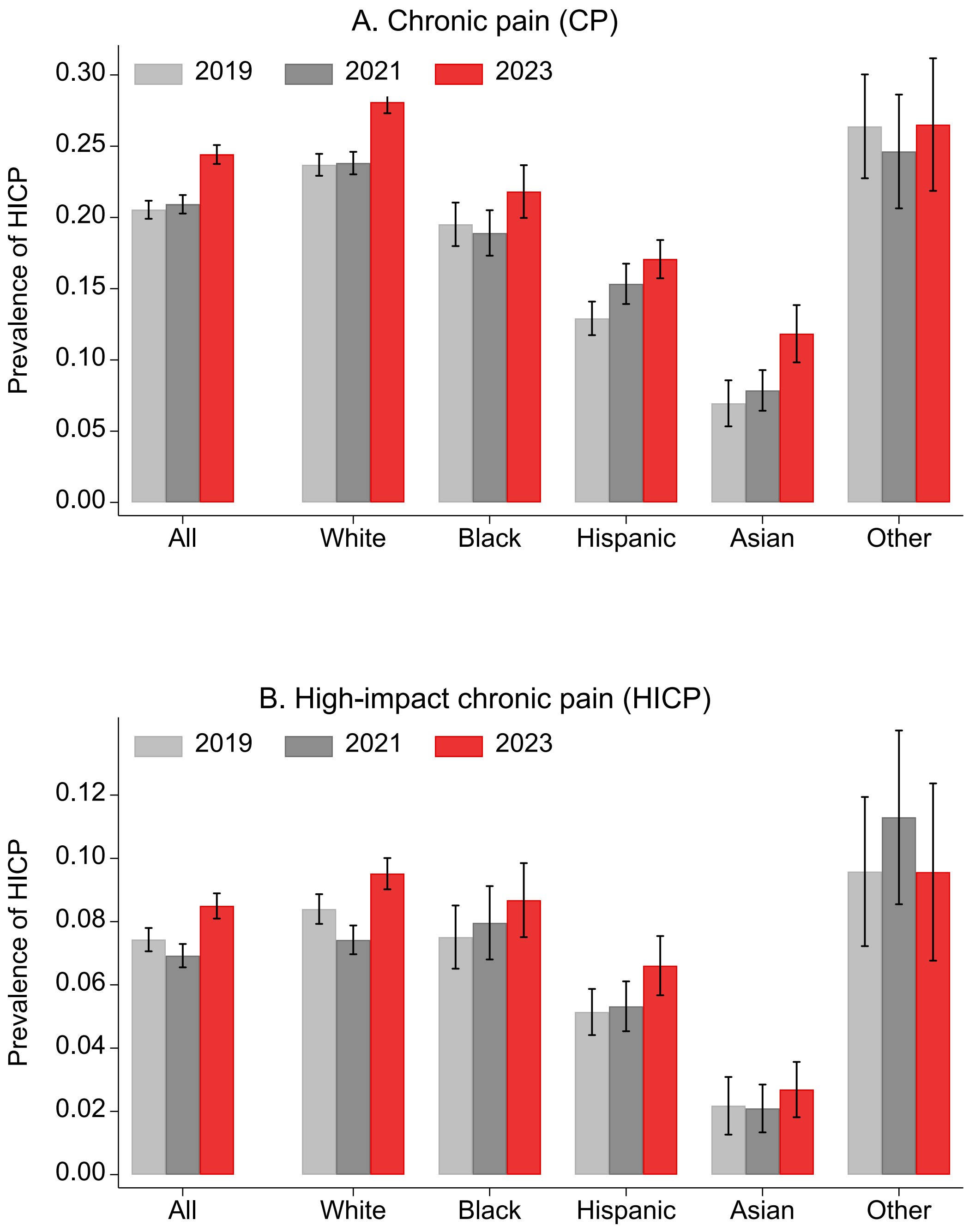

Note: Weighted prevalence estimates and 95% confidence intervals adjusted for complex sampling design.
